# Communicating cancer treatment with pictogram-based timeline visualizations

**DOI:** 10.1101/2024.06.04.24308420

**Authors:** Helena Klara Jambor, Julian Ketges, Anna Lea Otto, Malte von Bonin, Karolin Trautmann-Grill, Raphael Teipel, Jan Moritz Middeke, Maria Uhlig, Martin Eichler, Sebastian Pannasch, Martin Bornhäuser

## Abstract

**Objective:** This study evaluated the legibility, comprehension, and clinical usability of visual timelines for communicating cancer treatment paths. We examined how these visual aids enhance participants and patient understanding of their treatment plans.

**Materials and Methods:** The study included two online surveys and one in-person survey with hematology cancer patients. The online surveys involved 306 and 160 participants respectively, while the clinical evaluation included 30 patients (11 re-surveyed) and 24 medical doctors. Participants were assessed on their ability to understand treatment paths provided with audio information alone or with visual aids. The study also evaluated the comprehension of key treatment terms and the ability of patients to recall their cancer treatment paths.

**Results:** Visual representations effectively communicated treatment terms, with 7 out of 8 terms achieving over 85% transparency as pictograms, compared to 5 out of 8 for comics and 4 out of 8 for photos. Visual treatment timelines improved response quality, increased confidence, and were rated higher in information quality than audio-only information. In the clinical evaluation, patients showed good comprehension (mean response quality: 0.82) and recall (mean response quality: 0.71 after several weeks), and both patients and physicians found the visual aids helpful.

**Discussion:** We discuss that visual timelines enhance patient comprehension and confidence in cancer communication. We also discuss limitations of the online surveys and clinical evaluation. The importance of accessible visual aids in patient consultations is emphasized, with potential benefits for diverse patient populations.

**Conclusion:** Visual aids in the form of treatment timelines improve the legibility and comprehension of cancer treatment paths. Both patients and physicians support integrating these tools into cancer treatment communication.

## Introduction

The National Academy of Medicine/USA defines high-quality care as encompassing safety, effectiveness, timeliness, efficiency, patient-centeredness, and equity ^1^. Important for patient-centeredness and equity is an effective communication between health care providers and patients ^2–4^. Comprehensible information and patients’ health literacy, i.e. the ability to understand written and verbal medical information about diagnosis, prognosis, uncertainties and risks, are important in shared decision-making ^5^. However, mismatches in numeracy, literacy, and experience frequently challenge physicians communication with patients ^6^. Around 10% of the global population is estimated to lack basic literacy and, at a lower percentage, also numeracy skills, and even among those with high school education, adults have comprehension difficulties ^7–9^. Additionally, medical teams often encounter non-native speakers and patients with cognitive decline due to age or neurotoxic therapies, raising concerns about their understanding of treatment regimens for informed decision-making and further challenging the process ^5^.

Health literacy gaps are well-documented obstacles to equitability in care. Consent forms are frequently written in inaccessible language and illegible print ^3,10^. Likewise, verbal communication is often overly complex, with medical teams often overestimating patients’ literacy levels ^11–14^. This complexity is exacerbated when discussing intricate medical information, such as cancer treatments ^15–17^. Consequently, studies consistently find that patients tend to recall only half of their medical information ^17–22^, leading to implications for patients safety, treatment adherence and health outcomes ^12,23,24^.

Visual aids have proven to be beneficial for understanding the data, especially in the case of risks, uncertainties, and numerical information ^25–27^. In health care, visual aids are beneficial when promoting healthy choices to improving treatment adherence and risk-avoidance ^28–32^. Information that is supplemented by comics or pictograms measurably enhances health understanding and is perceived as helpful by patients ^30,33,34^. This approach is particularly helpful for vulnerable and non-native speaking patients, with whom visual aids are more effective even than translations ^30,33^. Despite these advantages, visual aids are at present underutilized in patient communication ^35,36^. The overall aim of this study was to develop and evaluate visual timelines for communicating cancer treatment paths using three haematological neoplasms as case studies.

## Results

In consultation with the patient board, the medical team, and based on feedback from our target audience ^6^, we had developed visual timelines for communicating cancer treatment paths (figure 1). To evaluate the effectiveness of these visual aids, we: (1) tested the clarity of the pictograms used; (2) assessed patients’ comprehension of treatment paths information with or without these visual timelines; and (3) tested their use in a clinical setting.

**Figure 1.**
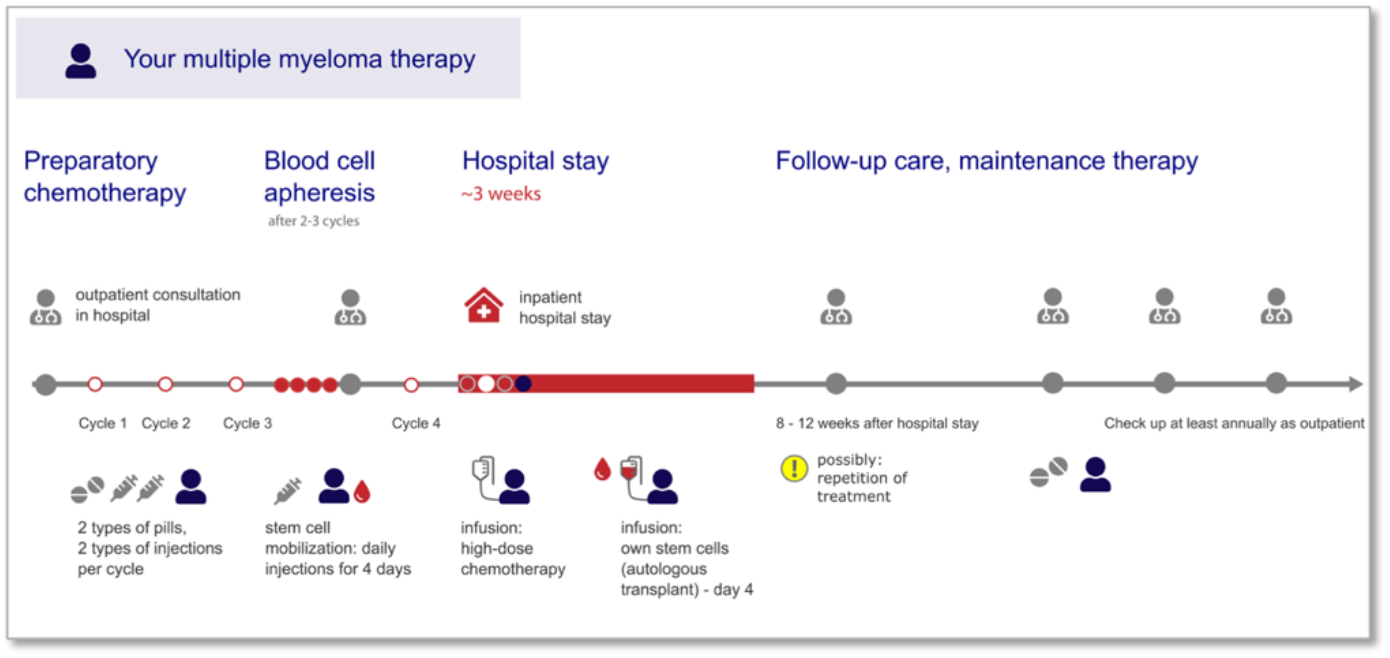
Example of a visual treatment timeline co-designed with patients and evaluated for comprehension with participants and patients. Multiple myeloma treatment with high-dose chemotherapy and autologous stem cell transplantation.

### Study 1 - Transparency and translucency of visual representations

We compared pictograms to comics and photos in their effectiveness to communicate medical term (figure 2A). We tested eight terms relevant for communicating cancer treatment paths (supplementary figure 1). Of the eight terms, six pictograms, five comic representations, and four photos were correctly identified (guessed) by at least 85% of participants and therefore met the American National Standards Institute (ANSI) transparency criterion of being understandable/guessable by at least 85% of participants (figure 3A, table 2A, B). Of the eight terms, six pictograms and comics, and five of eight photos were also rated as suitable by at least 85% of participants, and thus fulfilled the translucency criterion (figure 3B, Table1). Visual representations that did not pass the transparency or translucency criterion were the pictogram for “Pill” (76%), the comic for “Person” (77%), and the photos for “Hospital” (43%), “Person” (60%) and “Blood” (47%), which were neither guessable, nor considered suitable by >85% of participants, the ANSI requirements (85%) for symbols, and most are even below the somewhat more flexible standard of the International Organization for Standardization (ISO) of being understood by at least 67% of users without explanatory text ^47^. The visual representations for “infusion therapy”, arguably a highly specific term, was guessable by only 18% of the participants, but when prompted rated as “very suitable” in all visual representations (90-94%).

**Table 1.** Cohort descriptions.

| Feature | Category | Number | Percent % |
| --- | --- | --- | --- |
| **Study 1 Transparency and translucency of visual representations** |  |  |  |
| Age | <20 | 13 | 4 |
|  | 20-39 | 163 | 53 |
|  | 40-59 | 99 | 32 |
|  | 60+ | 31 | 10 |
| Gender | Male | 72 | 24 |
|  | Female | 228 | 75 |
|  | Diverse | 3 | 1 |
|  | Not answered | 3 | 1 |
| Health literacy<sup>51</sup> | High | 290 | 95 |
|  | Low | 16 | 5 |
| **Study 2 Information delivery formats for cancer treatment timelines** |  |  |  |
| Groups | Audio (a) | 60 | 38 |
|  | Pictogram (p) | 47 | 29 |
|  | Text (t) | 53 | 33 |
| Age | <18 | 1 (p:1,t:0,a:0) | <1 |
|  | 18-30 | 66 (p:18,t:26,a:22) | 41 |
|  | 31-60 | 85 (p:25,t:27,a:33) | 53 |
|  | 60+ | 7 (p:2,t:1,a:4) | 4 |
|  | Not answered | 1 (p:1,t:0,a:0) | <1 |
| Cancer knowledge | Large | 19 (p:5,t:7,a:7) | 12 |
|  | Some | 82 (p:23,t:34,a:25) | 51 |
|  | None | 58 (p:19,t:12,a:27) | 36 |
|  | Not answered | 1 (p:0,t:1,a:0) | <1 |
| **Study 3 Clinical evaluation of visual treatment timelines** |  |  |  |
| Gender | Male | 19 | 63 |
|  | Female | 11 | 36 |
| Age | 40-49 | 5 | 17 |
|  | 50-59 | 12 | 40 |
|  | 60-69 | 11 | 37 |
|  | > 70 | 2 | 7 |
| Disease entity | Multiple Myeloma | 21 | 70 |
|  | Lymphoma | 6 | 20 |
|  | AML | 3 | 10 |

**Table 2A.** Summary of transparency and translucency of visual representations. Visual representations that meet the American National Standards Institute (ANSI) requirement (recognizable to at least 85% of participants) are highlighted in bold, those below the threshold are shaded in grey. Included participants: 306

| Term | Transparency<br>(% correct) |  |  | Translucency<br>(% correct) |  |  |
| --- | --- | --- | --- | --- | --- | --- |
|  | Pictogram | Photo | Comic | Pictogram | Photo | Comic |
| MD | **90.2** | **90.6** | **90.6** | **95.4** | **96.4** | **85.7** |
| Hospital | 86.6 | 43.0 | 94.1 | 98.0 | 65.5 | 99.7 |
| Person | 87.9 | 60.6 | 77.5 | 94.5 | 73.3 | 73.0 |
| Infusion | 87.6 | 90.2 | 82.7 | 96.1 | 98.4 | 96.7 |
| Infusiontherapy | 18.6 | 28.3 | 33.9 | 92.8 | 94.1 | 90.2 |
| Pills | 76.2 | 95.8 | 86.3 | 81.1 | 99.1 | 85.0 |
| Syringe | 87.6 | 86.3 | 88.3 | 96.7 | 100 | 99.7 |
| Blood | 90.2 | 47.2 | 85.7 | 98.4 | 84.0 | 96.4 |

**Table 2B.** Statistical tests transparency of visual representations Included participants: 306.

| Test | Frequencies (n) | | | Cochrans Q Test<br>($\chi^2(2)$, p) | Pairwise Chi-square test, significance<br>level ($\chi^2(2)$, p) | | |
| --- | --- | --- | --- | --- | --- | --- | --- |
| Term | Picto | Photo | Comic | Across visual<br>representations | Picto : Photo | Picto : Comic | Photo:Comic |
| MD | 277 | 278 | 278 | 0.043, $p = .979$ | | | |
| Hospital | 266 | 132 | 289 | 224.66, $p < .001$ | -0.436,<br>$p \leq .001$ | 0.075,<br>$p = .126$ | 0.511,<br>$p \leq .001$ |
| Person | 270 | 186 | 238 | 83.60, $p < .001$ | 0.274,<br>$p \leq .001$ | 0.104,<br>$p < .05$ | -0.169,<br>$p \leq .001$ |
| Infusion | 269 | 277 | 254 | 14.35, $p < .001$ | -0.026,<br>$p = .583$ | 0.049,<br>$p < .05$ | 0.075,<br>$p \leq .001$ |
| Infusion-<br>therapy | 57 | 87 | 104 | 31.76, $p < .001$ | 0.098,<br>$p \leq .001$ | 0.153,<br>$p \leq .001$ | 0.055,<br>$p = .132$ |
| Pills | 234 | 294 | 265 | 53.46, $p < .001$ | -0.195,<br>$p \leq .001$ | -0.101,<br>$p \leq .001$ | 0.094,<br>$p \leq .001$ |
| Syringe | 269 | 265 | 271 | 1.10, $p = 0.578$ | | | |
| Blood | 277 | 145 | 263 | 195.93, $p < 0.00$ | 0.430,<br>$p \leq .001$ | 0.046,<br>$p = .530$ | -0.384,<br>$p \leq .001$ |

**Table 2C.** Summary of results from comparing information delivery formats for cancer treatment paths. Included participants: 160

| Variable | Audio with<br>Pictogram-based visual<br>aid | Audio with<br>Text-based<br>visual aid | Audio only | Significant difference between<br>groups, Tukey post-hoc |
| --- | --- | --- | --- | --- |
|  | Mean<br>(standard<br>deviation) | Mean<br>(standard<br>deviation) | Mean<br>(standard<br>deviation) | Mean difference (95% CI) |
| Response quality | 0.841<br>(0.117) | 0.820<br>(0.137) | 0.676 (0.163) | Picto:Audio: 0.12 (0.07, 0.20)<br>Text:Audio 0.16 (.10, 0.23) |
| Confidence rating | 0.78 (0.13) | 0.82 (0.11) | 0.55 (0.16) | Picto:Audio: 0.18 (0.13, 0.22)<br>Text:Audio 0.20 (.15, 0.24) |
| Rating of info quality | 0.79 (0.19) | 0.76 (0.21) | 0.62 (0.23) | Picto:Audio: 0.13 (0.13, 0.23)<br>Text:Audio 0.11 (.04, 0.18) |
| Total response time (sec) | 21.43 (9.22) | 21.71 (9.5) | 14.23 (5.76) | Picto:Audio: -7.21 (-10.85, -3.56)<br>Text:Audio -7.64 (-11.61, -4.12) |
| Total response time confidence (sec) | 4.19 (1.27) | 4.82 (2.06) | 5.14 (0.90) | No statistical difference (Welch- ANOVA) |

**Table 2D.** Summary of results from clinical evaluation of visual treatment timelines.

| Questionnaire | Participants | Response quality to content questions |  |  |  |
| --- | --- | --- | --- | --- | --- |
| Timepoint | n | Mean | Standard deviation | Median | Standard error |
| Survey (timepoint 1) | 30 | 0.82 | 0.15 | 0.8 | 0.03 |
| Re-survey (timepoint 2) | 11 | 0.71 | 0.23 | 0.71 | 0.07 |

**Table 2E.** Individual patients’ response quality over time.

| Patient | Response quality, survey | Response quality, re-survey | Change in response quality over time |
| --- | --- | --- | --- |
| ID | Mean | Mean | Delta |
| 13 | 1.0 | 0.6 | -0.4 |
| 14 | 0.8 | 0.8 | 0.0 |
| 15 | 0.6 | 0.6 | 0.0 |
| 16 | 1.0 | 0.8 | -0.2 |
| 17 | 1.0 | 0.4 | -0.6 |
| 19 | 0.8 | 1.0 | 0.2 |
| 20 | 1.0 | 1.0 | 0.0 |
| 21 | 0.8 | 0.6 | -0.2 |
| 28 | 1.0 | 1.0 | 0.0 |
| 26 | 0.4 | 0.4 | 0.0 |
| 27 | 0.8 | 0.6 | -0.2 |

**Figure 2.**
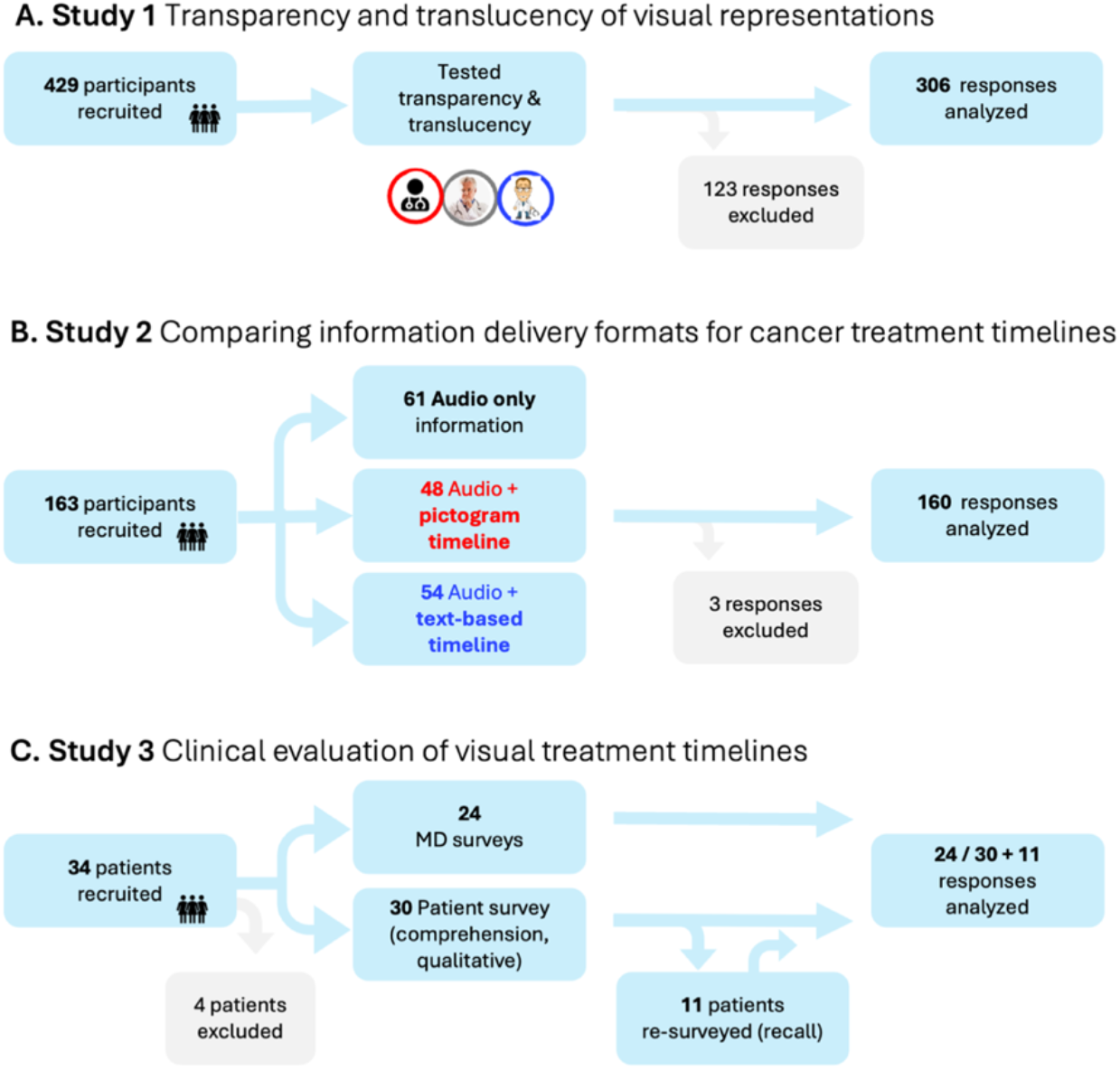
Flowcharts of results from the three studies.

**Figure 3.**
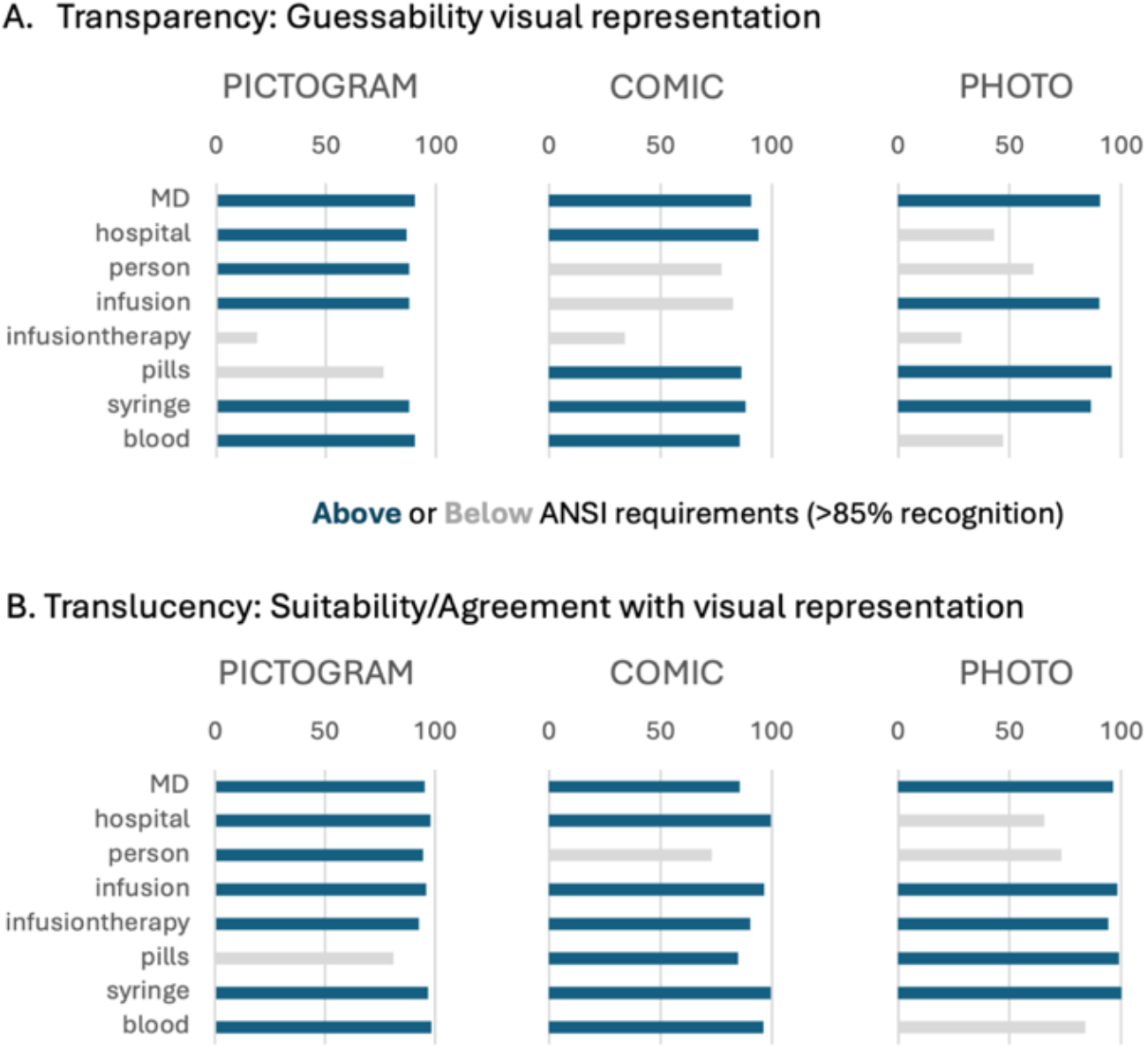
Transparency and translucency of visual representations. **A.** % Transparency: can participants guess the image term? Overall, many are above the 85% mark. Highest number of icons above 85% are pictograms, then comics, lowest photos. **B**. % Translucency: do participants rate the icon as suitable for a known term? Overall, many are above the 85% mark. Highest number of icons above 85% are pictograms, then comics, lowest photos. Participants: 306.

This data indicates a slight skew towards pictograms and comics being more guessable and suitable, however based on our eight tested terms, no visual representation was consistently outperforming the other. “MD” and “Syringe” were equally guessable in all forms of visual representation (Chi-squared <0 / 1.1, no deviation across all visual representations, table 2B), while infusion therapy was not sufficiently guessable in any representation (table 2A). For “Hospital”, “Person”, “Infusion”, and “Blood”, pictograms were significantly more guessable than comics and/or photos as conformed by Chi-squared testing (table 2B). Only for “Pill”, comic and photo representations significantly outperformed the pictogram (table 2B). A similar result was obtained for the suitability of visual representations. Again, for “Hospital”, “Person”, and “Blood”, pictograms were rated significantly more suitable than comics and/or photos, however not only “MD” and “Syringe”, but also “Infusion” and “Infusion therapy” were rated equally suitable in all visual representations.

Applying a validated health literacy test 51 revealed a relatively homogeneous cohort, with over 95% of participants demonstrating high health literacy (cohort description: table 1). As a result, we could not test differences in visual representation transparency and translucency across literacy levels. Also, as the group sizes across these age categories were not equal, testing for age-depended could would not reliably reveal effects. We did however observe that elderly participants were generally slower in their responses across all forms of visual representations, and that both younger and older participants responded fastest with pictograms than with photo representations (supplementary table 2).

### Study 2 – Comparing information delivery formats for cancer treatment timelines

We compared participants’ ability to understand cancer treatment path information presented as audio-only or with two different formats of visual treatment timelines (figure 2B, figure 1, supplementary figure 2, summary of results: supplementary table 4). Compared to participants who received only audio information, simulating a typical patient consultation, those who also received treatment timelines demonstrated significantly higher overall response quality (suggesting questions were easier to answer) when answering content question (figure 4A, 0.84/0.82 compared to audio 0.68). The response quality was not statistically different between participants receiving text-based and pictogram-based treatment timelines (table 2C). Participants with treatment timelines not only were quantitatively better in answering content questions, but also subjectively indicated feeling more confident their answers were correct (figure 4B, 0.78/0.82 compared to audio 0.55) and also rated the quality of the information higher (figure 4D, 0.79/0.76 compared to audio 0.62). Overall, the groups with treatment timelines were significantly slower in their response times than participants with audio information only (figure 4C, table 2C, supplementary table 2, 21.4/21.7 seconds compare to audio 14.2 seconds). Although the groups were slower in answering content-related questions, in comparison the time they required to rate their confidence in their answers was instead comparable across all three groups and not statistically different (figure 4E, 4.1/4.8/5.1 seconds). Thus, the slower response time may indicate that participants indeed make use of the visual aids when answering questions. Despite this slight delay, the groups with visual treatment timelines, both text and pictogram-based, showed significantly higher overall response quality, as well as for a higher question-level success ratio (figure 4F).

**Figure 4.**
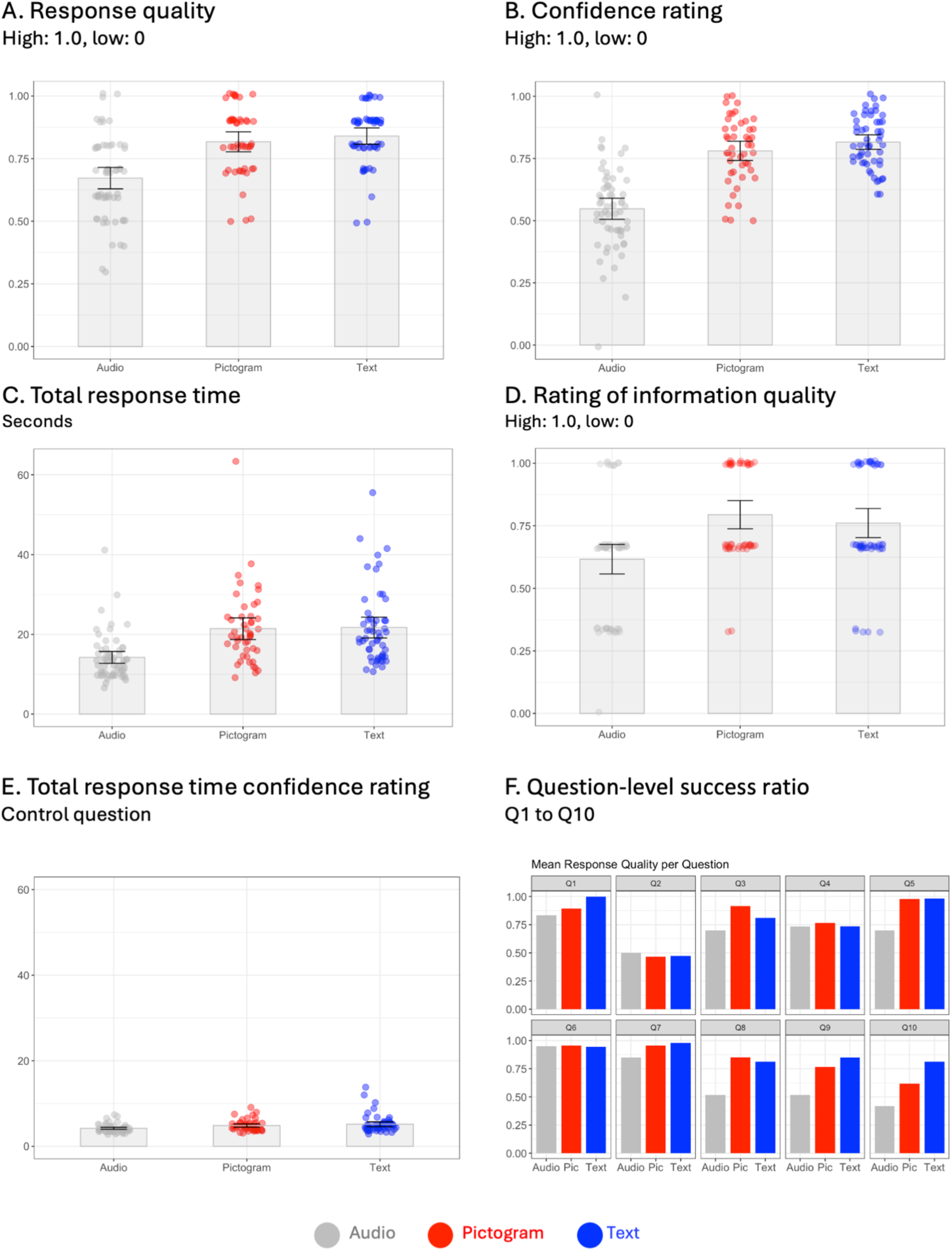
Comparing information delivery formats for cancer treatment paths. Visual aids (pictogram- and text-based) improved: **A**. the overall response quality with a higher value indicating that a question was easier to be answered correctly by participants, **B**. increased respondents confidence rating and **C**. response times, and **D**. were rated higher in information clarity. **E**. Response times for content questions varied, while the times for the control questions were similar across groups. **F**. Question-level success ratio indicate how easy the individual questions 1-10 (supplementary table 3) were answerable by participants in the three groups. Participants: 160.

### Study 3 – Clinical evaluation of visual treatment timelines

Given that visual timelines improved comprehension of cancer treatment paths, and that visual elements were clear to the majority of participants, including the relevant age group for haematological diseases, we next evaluated the effectiveness of visual aids in the clinic for three use-cases (figure 2C): patients treated for multiple myeloma with autologous stem cell transplantation (figure 1), patients undergoing allogeneic stem cell transplantation, and patients receiving CAR-T cell therapy (supplementary figure 3). We tested the visual treatment timelines in patient interviews and questioned attending MDs and patients. All MDs (n=24) fully or partially agreed that patients seem to understand the aids, and that aids were a helpful addition; almost all MDs partially or fully agreed that they were able to use visual aids without preparation and indicate that they would include aids in future communication (figure 5A). The 30 patients we surveyed (aged 44 to 72, average 58) were similarly positive, all responded that the aids helped during the interview and for answering questionnaire, and they plan to consult them again (figure 5B). For the five content questions the mean response quality was 0.82 (sd 0.15) and 5 of the 30 patients answering all questions correctly (figure 6A, table 2D).

**Figure 5.**
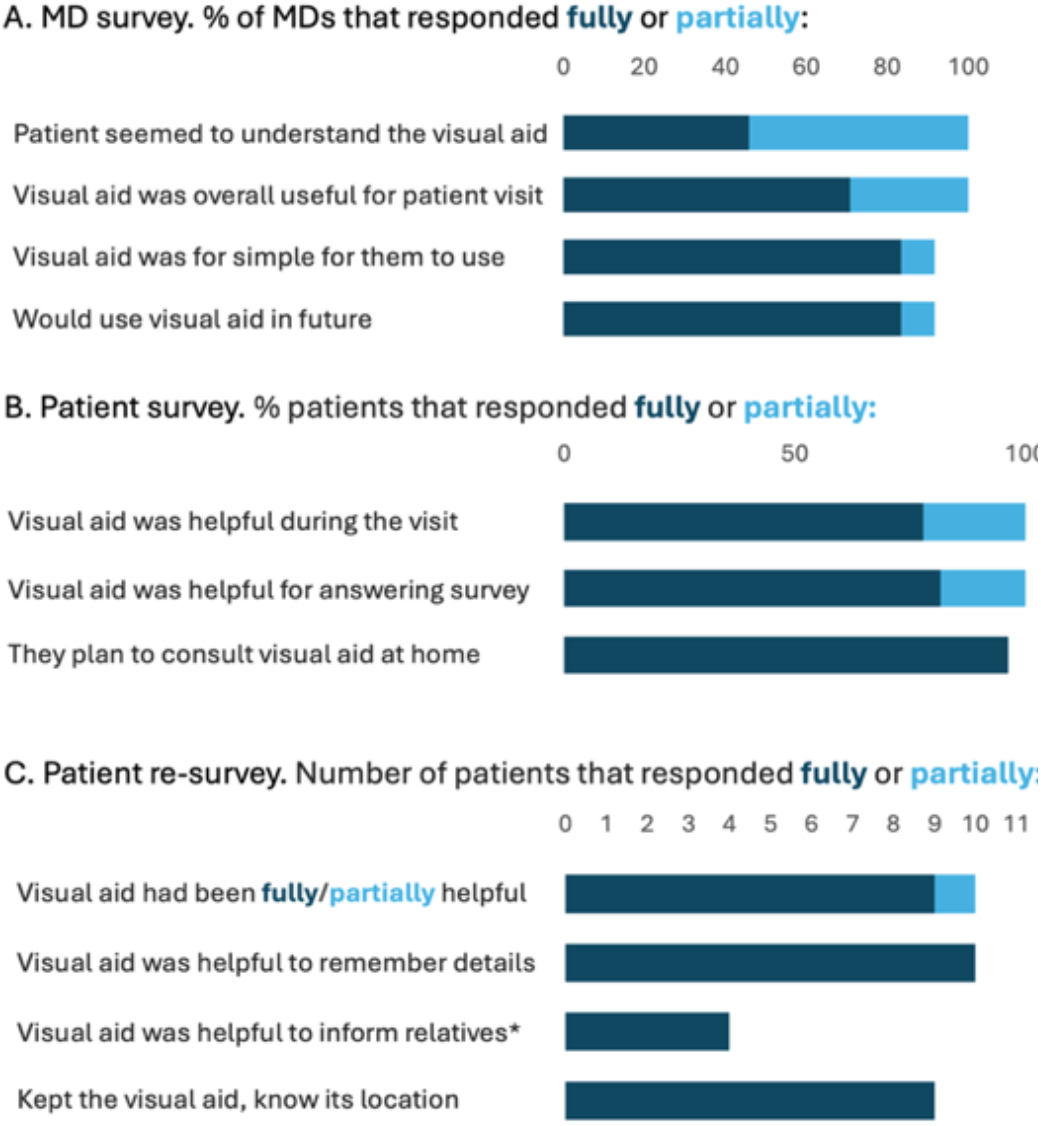
Clinical evaluation of visual treatment timelines. **A.** Responses from MDs that used visual treatment timelines for treatment paths in patient interviews, n=24. **B**. Responses from patients after interview with treatment plan, n= 30. **C**. Responses from patients at re-survey several weeks after initial interview, n=11. * missing responses: patients had not spoken with any relatives about their treatment.

**Figure 6.**
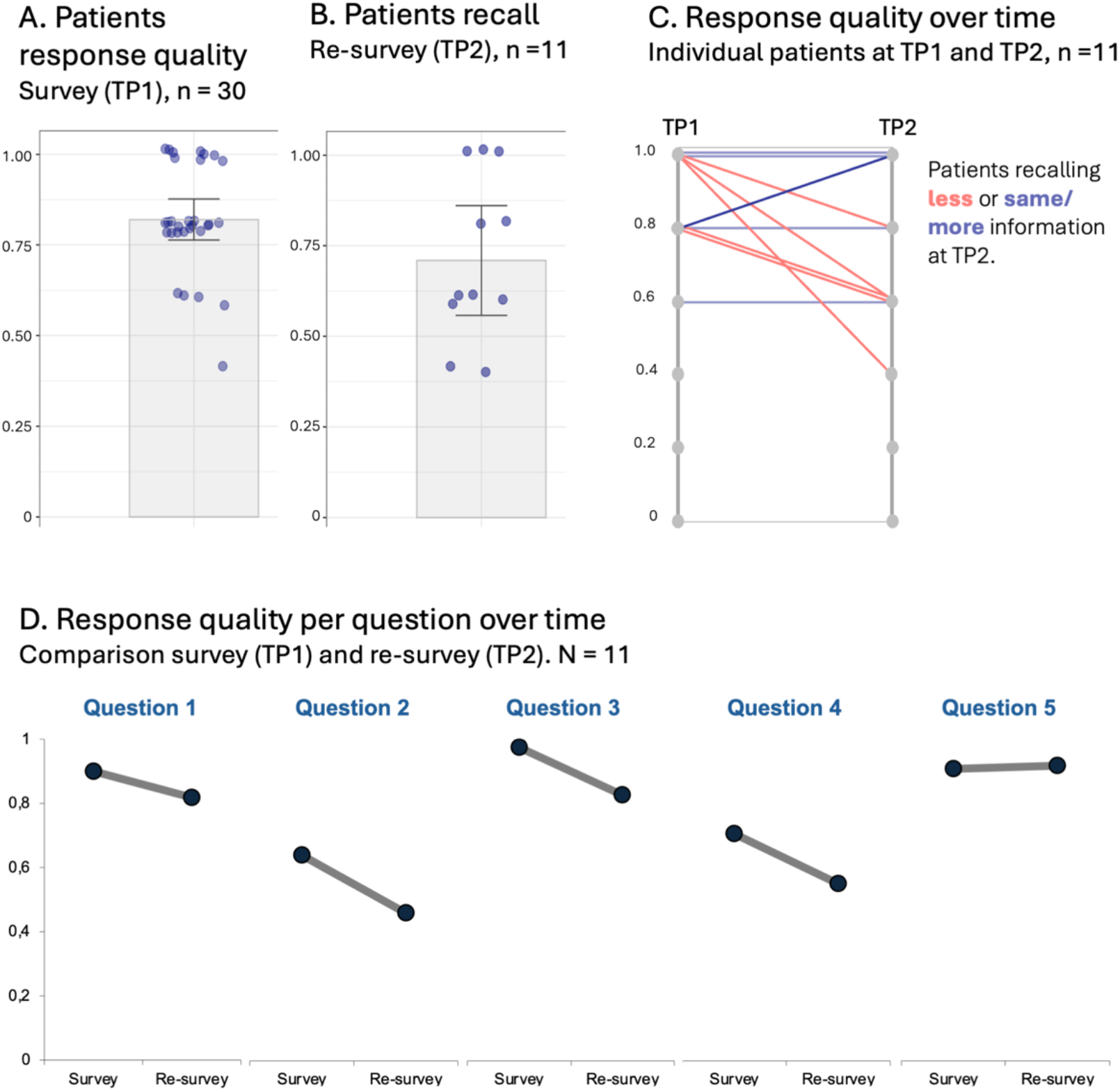
Clinical evaluation of visual treatment timelines. **A.** Patient response quality at outpatient consultation, n= 30. **B**. Patient recall response quality at re-survey when visiting for cell apheresis, n=11. **C**. Comparison of individual patients’ responses at survey and re-survey, n=11. **D**. Question-level success ratio at survey and re-survey, n=11. Questions: see supplementary table 3.

Patients with multiple myeloma return to hospital several weeks after the begin of therapy to undergo stem cell apheresis for the following autologous stem cell transplantation. At this point we were able to re-survey 11 of the initially 30 patients. While we observed a drop in overall response quality to 0.71 (sd 0.23, figure 6B), this is still a high recall rate, and 5 of the 11 patients remembered the same amount as right after the interview (figure 6C, table 2D, E). Some questions were easier to answer than others, we therefore also analysed the question-level success ratio at both survey time points. This revealed that while we saw a drop in response quality for each question (supplementary table 3), the overall response quality was still high after several weeks, with the question with lowest response quality still correctly answered by >50% of patients (figure 6D). Patients had kept their visual treatment timeline, and still fully or partially agreed that it had helped them understand the procedure. Moreover, they indicated that they had consulted the plan at home, and, if the spoke with relatives about their treatment (4/11), used it to refresh their memory, and even send pictures/photocopies of the plan, to relatives (figure 5C).

## Discussion

In this work, we investigated to what extent visual treatment timelines communicating the treatment path can effectively supplement health care information. We used visual treatment timelines for three haematology treatments as example cases. Survey results reveal that visual treatment timelines significantly enhance comprehension and increase participants’ confidence when responding to content questions on treatment paths. Consistent with existing literature ^31,41^, pictograms and comics often outperformed photo representations and were deemed suitable across various age groups, highlighting their accessibility and versatility in patient communication. Our data also show that some visual representations were not sufficiently guessable, therefore, legibility should be evaluated for each visual representation and, when used, pictograms should be combined with an explanation and a legend.

In our clinical evaluation, MDs and patients positively responded to integrating visual treatment timelines in consultations, and patients remembered treatment details to correctly answer questions immediately as well as several weeks after the interview. Patient consultations take place under time pressure as staff is obliged to provide comprehensive and legally compliant information on various aspects of treatment. The American Cancer Society recommends that patients request decision aids, e.g. in the form of written treatment plans or schedules 37. Thus, visual aids like our treatment timelines, designed with minimal text and supplemented with pictograms, could effectively complement patient interviews.

Interestingly, we found that the pictogram-based visualization and the text-based treatment timeline were equally helpful for recall among study participants, suggesting that any form of supportive information is better than none ^27^. Research on multimedia learning supports this, indicating that a mixed-format approach is more effective than relying on a single channel ^52^. From an information design perspective, the text-based timeline, despite lacking decorative elements like pictograms or colour, qualifies as a visual aid due to its organized layout along an axis ^53^. However, effectiveness of specific visual aids is depended on the communication goals, which could range from enhancing recall of information to influencing perceptions or encouraging specific health behaviors ^48^. Further research is needed to determine whether a purely text-based description can be just as effective for recall and trust and to understand how readers perceive and differentiate these two forms of visual information.

Currently, visual aids are underutilized in patient consultations ^35^. Although physicians acknowledge the value of visual aids, they are rarely used, mainly because visual aids are not readily available. Our survey of existing patient information materials revealed that information figures and data visualizations are largely missing in brochures for cancer treatment. This aligns with findings from a previous systematic review ^36^. In aging societies, where the number of elderly cancer patients is still rising, effective healthcare and risk communication present a significant challenge. This challenge could be mitigated by using appropriate visual aids for patients that help the various communication goals in healthcare, from identifying and recalling information, to behavioural adjustments ^48,54–56^.

A limitation of our study is the surveyed demographics in the clinical evaluation, as only a small number of patients were available locally during the recruitment period. The limited number of patients also meant that in this first clinical evaluation we could not randomize patients into two arms, a control and an experimental group. However, based on our initial evaluation conducted with the target audiences ^6,47^, and with feedback from the patient board, we conclude that the visual cancer treatment timelines do significantly enhance comprehension within our target group. A logical next step therefore is a multi-centric, controlled clinical trial to pave the way for clinical adoption. In an aging society haematological neoplasms among elderly is still rising ^57^ and treatments become more complex with advancements in patient stratification and personalized medicine ^58,59^, making accessible patient information even more pressing. A trial could also compare several realizations of the visual treatment plan and possibly also test measurable effects on the quality of life.

Our visual treatment timelines likely have broader applications, e.g. for other cancer types or other long treatment schedules. Such visual aids may also fill the information need of the elderly patients, experiencing an anxiety-inducing diagnosis ^16,23,60–62^ and could generally support vulnerable populations, children patients, relatives and caregivers, nurses ^6^. Given that the online surveys (Study 1-2) were skewed towards a younger population with high health literacy (Study 1) or prior knowledge of cancer treatment (Study 2), our data provides limited insights into vulnerable populations. It is possible that group-based differences, such as variations in response time or confidence, could manifest more subtly. However, the granularity of our study does not allow for a detailed analysis of these potential effects.

The rapid developments of AI-based tools likely will also facilitate generating visual aids from text-prompts. Several resources are already available that offer a wide range of health-related icons/pictograms, which could enhance the visual treatment plans. Examples are Smart Servier, BioIcons, SVGrepo, and Health Icons. Health Icons (healthicons.org/) provides access to over 1,300 medical icons for anatomical, disease-related and treatment-related terms and medical devices under a public domain/CC0 license. These icons can be easily integrated into software tools like interactive dashboards or AI-based applications, expanding their utility in healthcare settings. At times, modest measures can have profound effects, as was demonstrated by improved cancer survival when monitoring patients well-being with questionnaires ^63^. Patients expressed gratitude for these visual aids, treasuring them as they navigate their health care journey. The aids provide tangible answers to important questions that were also raised by the patients involved in this study, such as “How long will I be away from home?” and “How often do I come back to the hospital?”.

## Funding statement

This work was supported by an habilitation award of the Medical Faculty Carl Gustav Carus of the Technische Universität Dresden, Germany to HKJ; project funding of the Mildred Scheel Early Career Center Dresden P2/German Cancer Aid (Deutsche Krebshilfe, project ID: 74MSNZNGAr); project funding of the Hochschulstiftung Medizin Dresden, Germany.

The funders had no role in study design, data collection and analysis, decision to publish, or preparation of the manuscript.

## Competing Interests Statement

The authors have no competing interests associated with this work.

## Author Contributor ship Statement

HKJ, SP, MB were jointly involved in the project conceptualization, methodology, supervision and writing of the manuscript. HKJ, JK, ALO survey conceptualization, methodology and data analysis. HKJ and JK data visualization. HKJ and MB Funding Acquisition. All authors were involved in the investigation, validation, reviewing and editing of the manuscript.

## Acknowledgement

We thank the anonymous respondents of our survey and the medical doctors and nurse staff at the Medical Clinic 1/University hospital Dresden for participating in the clinical evaluation. We thank the team of the transfusion medicine (head Kristina Hölig) for scheduling the re-surveys. We thank Ava Elise Lehrach for help with digitizing questionnaire data. We thank the patient board of the NCT-UCC Dresden for feedback and support.

## Data availability statement

The data, study materials, and supplementary data are freely available at our Open Science Framework https://doi.org/10.17605/OSF.IO/WKQB4. The protocol for study 1 (Transparency and translucency of visual representations) was preregistered: https://osf.io/cs57n; the protocol for Study 2 (Information delivery formats for cancer treatment timelines) was preregistered: https://osf.io/t2gkq).

## Methods

To develop visual aids in the form of visual treatment paths for patients with haematological neoplasm (figure 1, supplementary figure 3), we assessed the information needs by observing outpatient consultations as well as via meetings with patients, patient board, and clinicians, and guides for patient treatment plans and schedules ^37^. This revealed the time-course of treatment, the sequence of interventions, and their settings (hospital stay or outpatient care) as relevant areas for visual aids. In existing public information from national cancer institutes and cancer charities in the USA, UK, and Germany treatment timeline visualizations ^38^ were scarce in the text-heavy brochures, and mostly limited to photos and anatomical illustrations (supplementary table 1). Only 3 of the 44 figures provided some information on the treatment timelines.

Using an iterative design approach ^39–42^ that included the target group ^6^, we then developed visual treatment timelines, the final version of which was used in this study. In this work, we evaluate the usability of the visual treatment timelines. In the first step, Study 1 compared the legibility of different visual representations for key terms. Next, Study 2 was used to assess participants’ comprehension when receiving information either through audio alone or supplemented with text- or pictogram-based visual treatment timelines. Studies 1 and 2 were conducted with participants in an anonymous online survey and not with patients. Finally, Study 3 evaluated the clinical use of visual treatment timelines with patients.

### Study 1: Design and Evaluation of Visual Representations

#### Design

Essential for designing a visual aid is the identification of suitable visual representations to encode the key terms. We have selected pictograms for the visual treatment timelines as they are widely used e.g., in public transport ^41^, have been integrated in health information ^31,43,44^, and are highly rated by patients ^34,45,46^. However, given their high abstraction level, pictograms must be evaluated before use with the target audience ^47^. By ANSI (American National Standards Institute) requirements, only visuals that are recognizable by at least 85% of participants fulfil the criteria for being self-explanatory and helpful ^47^. Alternative visual representations are photographs and comics, but these may contain irrelevant information, e.g., gender of medical professional, or overemphasize details (comics).

To compare visual representations, we designed a one-factorial (2 phases) within-subjects design (figure 2A). In each phase, eight terms (supplementary figure 1) were shown in three different visual representations (pictogram, comic, photo). To minimize the order effect, the visual representations were shown in random order. The participants were required to answer questions on the transparency (phase 1) and translucency (phase 2) of the visual representations. To assess transparency (guessability, Question: *“What is the meaning of the prompted visual?”*), participants were required to enter free text to describe a visual representation with one term. This was matched to a syntax of correct terms. To examine translucency (Question: *“Is the prompted visual suitable for term?”*), participants assessed the suitability of the visual representation and its term on a scale from 1-7 (1-4: not appropriate; 5-7: appropriate). The study was preregistered at OSF (https://osf.io/cs57n).

#### Participants and Power Analysis, Procedure

Inclusion criteria were ability to understand, read and write German. No personal data was collected so that the identity of the participants was completely protected. Since no identifiable personal information was obtained, this survey did not fall under the requirements for ethical review board approval at the TU Dresden. We targeted a sample size of 259 based on a power analysis with a desired power of 0.8, alpha level of 0.05, and an assumed medium effect size of 0.1. Based on inclusion/exclusion criteria, we included 306 participants (mean age 39) in the study, see table 1 for cohort description.

The online, open-label cohort questionnaires were conducted in German, administered using LimeSurvey software, and piloted to validate questions and solve technical issues. Online participants were recruited via social media, notice boards, and university mailing-lists. Participants had to provide informed consent, agree to anonymous responses being used for research, were provided with contact information of researchers, and the opportunity to withdraw.

#### Statistical Analysis and Data Visualization

Data on transparency and translucency were analyzed using SPSS Version 28.0.0.0. Transparency was assessed by the frequency of correct free-text descriptions, and translucency by the suitability ratings. To test the comprehensibility of the visual representations, their transparency (guessability, participants had to add a free text description of the image) was matched to a corpus/syntax of all correct descriptors; terms not in syntax were considered incorrect. We then summarized the frequency of correct answers. To assess the translucency of the visual representations (question: ‘Is the prompted visual suitable for the term?’), the participants’ ratings of visual representations were assigned a numerical value (step 1 to 4, not appropriate/0, steps 5-7 appropriate/1) and then summarized by frequency for each term.

### Study 2: Comparison of Information Delivery Formats for Cancer Treatment Timelines

#### Design

To compare the effectiveness of three formats for delivering information on cancer treatment paths, we quantified participants’ response qualities using a between-subject design with multiple comparisons (figure 2B). Participants were randomly assigned to one of the three groups, each corresponding to a different treatment condition (the primary independent variable): (i) audio only (scenario in current patient consultation without reading materials), (ii) audio with pictogram-based treatment timeline (supplementary figure 2A), or (iii) audio with text-based treatment timeline (supplementary figure 2B). Group (ii) and (iii) received time course data as flow-chart/timeline and they were tasked to identify specific information with access to the stimulus and to rate their trust/confidence ^48^. All participants received some general orienting information about leukaemia before starting the survey. All participants then received the 2-minute audio information about treatment and timelines. Participants then answered 10 content questions (multiple choice, see supplementary table 3). After each content question, participants were asked to rate their confidence in answering (4-step Linkert rating scale, “How confident are you in your answer?”, answers: unsure, somewhat unsure, somewhat sure, sure). Participants in group (ii) and (iii) could use the respective treatment timelines while answering questions.

After the completion, participants were asked to rate the quality of the received information using a 4-step Linkert scale (answers: incomprehensible, rather incomprehensible, rather understandable, understandable). We included a question on prior health education to monitor a potential selection bias (self-assessment of prior knowledge on cancer, answers: no/some/extensive prior knowledge). The study was preregistered at OSF (https://osf.io/t2gkq). The timing of each step was recorded.

#### Participants and Power Analysis, Procedure

The inclusion criteria were identical to that of Study 1. To obtain reliable differences in responses, we set the desired statistical power at 0.8 and chose an alpha level of 0.05 and, given the lack of previous studies, assumed a medium effect size of 0.25, which revealed a required sample size of 159. 160 participants were included in the analysis of the effect of visual treatment timelines (mean age 38, table 1). In the comparison of visual representations, 306 participants were included with a mean age of 39 (cohort description see table 1). The same software and the same procedure were used as in Study 1.

#### Statistical Analysis and Data Visualization

Data were analyzed using SPSS Version 28.0.0.0. The effectiveness of the information delivery formats was evaluated by calculating several key parameters. The participants’ response quality was calculated as the ratio of the correct answers of the participant to all 10 multiple-choice questions. For the total response times (content question, confidence/control questions) the time was summed up. For confidence rating, Linkert scale responses were assigned a corresponding numerical value (1 - unsure, 2 - somewhat unsure, 3 - somewhat sure, 4 - sure), then averaged per participant across all answers and normalized to a continuous 0 (low) to 1 (highest) scale to be comparable in scale to the other observations. For rating of information quality, the Likert scale responses were assigned a corresponding numerical value (1 - incomprehensible, 2 - rather incomprehensible, 3 - rather understandable, 4 - understandable) and then transformed to a 0 (lowest) to 1 (highest) scale to be comparable in scale to the other observations (0 - lowest rating, 0.33 - slightly positive rating, 0.67 - moderately positive rating, 1 highest rating). The question-level success ratio was calculated as the ratio of correct participants’ answers to all participants, indicating how well the question was answered across all participants.

A one-way ANOVA was employed to compare the means of the three groups to test for statistically significant group effects, and variance homogeneity was confirmed with Levene’s test. For those cases where the ANOVA indicated a significant difference, we tested the homogeneity of variances using Levene’s test. If Levene’s test indicated a difference in variances, we then performed a Tukey post hoc test to determine which specific group differences were statistically significant. When ANOVA indicated a significant group difference, a Tukey post hoc test was employed to identify specifically which groups are different from each other.

### Study 3: Clinical Evaluation of Visual Treatment Timelines

#### Design

We evaluated the clinal usability of visual treatment timelines for cancer treatment timeline communication with a non-blinded, open-label patient questionnaire (figure 2C). The questionnaire included five multiple-choice content questions (a subset of the questions similar to those used in Study 2, see supplementary table 3), three questions with rating scales (5-step Likert scale, usefulness of the visual aid, answers: very/somewhat/not helpful, distracting, not sure), and one open-ended question (*“How do you envision to/did you use the visual treatment timelines at home?”*). The questionnaire was piloted and validated with medical doctors (MDs) and patient members of the patient board of the National Center for Tumor Diseases/NCT-Dresden. The clinical evaluation, including the patients consent form, was approved by the TU Dresden ethics board (BO-EK-338072022). Patients received time course data as flow-chart/timeline and were tasked to recall and rate the information ^48^.

#### Participants

Inclusion criteria were diagnosis of a hematologic neoplasm, age ≥ 18 years at diagnosis, attended at the Medical Clinic 1, University Hospital Dresden, ability to give consent; exclusion criteria were inability to complete a structured questionnaire, e.g., in the presence of comorbid dementia, insufficient language skills, illiteracy. Patients were informed about the purpose and design of the study as well as familiarized with visual treatment timelines for treatment path communication during the consultation. After giving consent, patients were informed by the MD about the treatment procedure with the visual treatment timeline (figure 1, supplementary figure 3) before answering the questionnaire. No statistical tests were planned for the clinical evaluation with patients; thus, no power analysis was required. In total 34 patients were recruited, of which 30 were included (cohort description see table 1) along with 24 questionnaires by their attending MDs. Patients treated for multiple myeloma were additionally invited to the re-survey when returning for a scheduled stem cell apheresis. All data was handled in compliance with the TU Dresden ethics board.

#### Procedure

Patients completed a paper-based questionnaire after being informed about their treatment procedure with the visual timeline. The initial questionnaire was administered in-person, and no additional information was provided before the re-survey. For the re-survey, patients were not provided with any additional verbal, written, or visual information and were interviewed before consultation with a medical doctor.

#### Statistical Analysis and Data Visualization

For the clinical evaluation, descriptive statistics were used to summarize the frequency of responses. All data plots were prepared using R and ggplot2, version 4.3.2 ^49,50^.

### Patient and Public Involvement

Our work was supported by the patient board of the National Center for Tumor Diseases/NCT-Dresden, which also includes former patients. The planned work and its progress were presented to the entire board. A project advisory group of three board members was also involved in reviewing and piloting the questionnaire and provided helpful input on the design of visual treatment timelines. The ongoing project was presented publicly at “patient day’s” organized by the National Center for Tumor Diseases.

## Supplementary Materials

*Supplementary Figure 1. Visual representations compared for transparency and translucency (Study 1) and Supplementary Figure 2. Text-based visual aid used for comparing information delivery formats (Study 2): see OSF repository.*

**Supplementary Figure 3.**
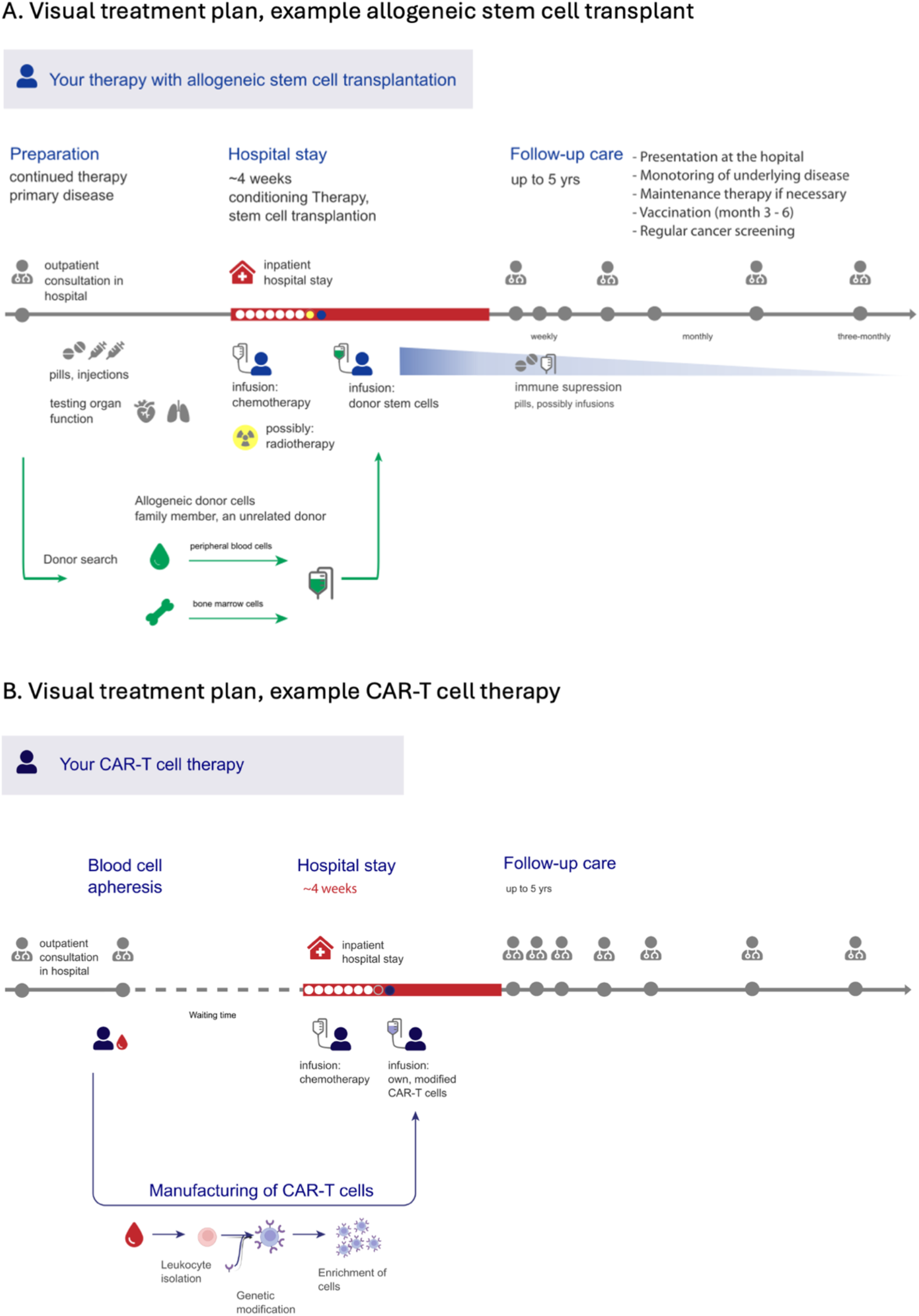
Additional visual aids used for the clinical evaluation (Study 3)

**Supplementary Table 1.** Existing patient information. Comparison of public information for three haematological neoplasm therapies (MM, multiple myeloma; AlloTX, allogenic transplantation for e.g. AML patients; CAR-T, targeted immunotherapy for e.g. lymphoma patients) in USA, UK and Germany (DE). The information includes text only or few visuals with decorative character (figure of cell types, patient). In three cases a process was visualized, each without details.

| Organization (Country) | Case | Text | Visual elements<br>(photos, diagrams) | Visual of<br>process |
| --- | --- | --- | --- | --- |
| NHS (UK) | MM | Website | - | - |
| Myeloma UK (UK) | MM | 12-page PDF | - | - |
| Myeloma UK (UK) | MM | 58-page PDF | 3 | - |
| Cancer research UK | MM | Website | 2 | - |
| Cancer research UK | MM | Website | 2 | 1 |
| NCI (USA) | MM | Website | 4 | - |
| KID (DE) | MM | Website | 1 | - |
| Onkopedia (DE) | MM | Website | 4 | - |
| Myelom-net (DE) | MM | 60-page PDF | 9 | - |
| Myelom.org (DE) | MM | Website | - | - |
| Lymphome-de (DE) | MM | 12-page PDF | 2 | - |
| DLH (DE) | MM | 50-page PDF | 4 | - |
| NHS (UK) | AlloTX | Website | - | - |
| Cancer research UK | AlloTX | Website | 2 | 1 |
| NCI (USA) | AlloTX | Website |  | 1 |
| KID (DE) | AlloTX | Website | 1 | - |
| Onkopedia (DE) | AlloTX | Website | - | - |
| NCI (USA) | CAR-T | Website | 2 | - |
| NHS England (UK) | CAR-T | 29-page PDF | - | - |
| KID (DE) | CAR-T | 2-page PDF | 1 | - |
| Lymphome-de (DE) | CAR-T | 12-page PDF | 4 | - |

**Supplementary Table 2.**
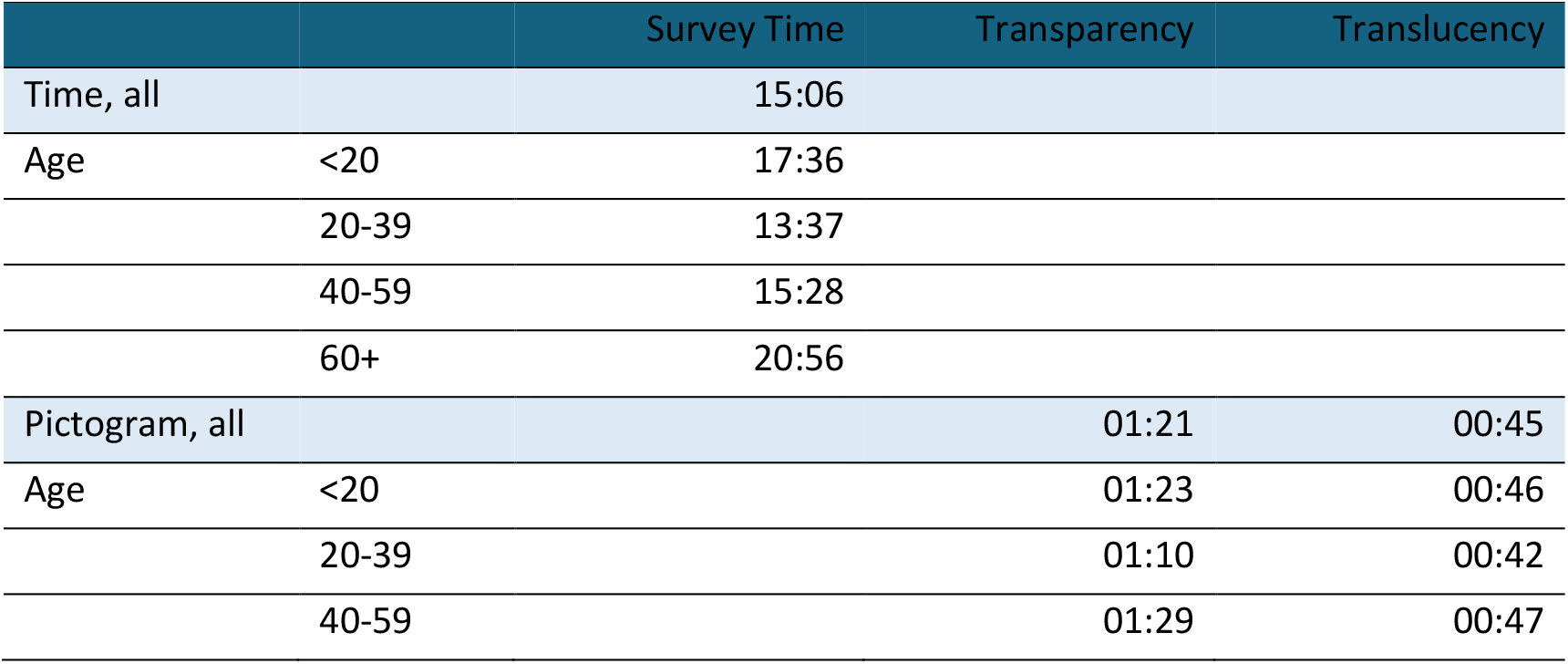
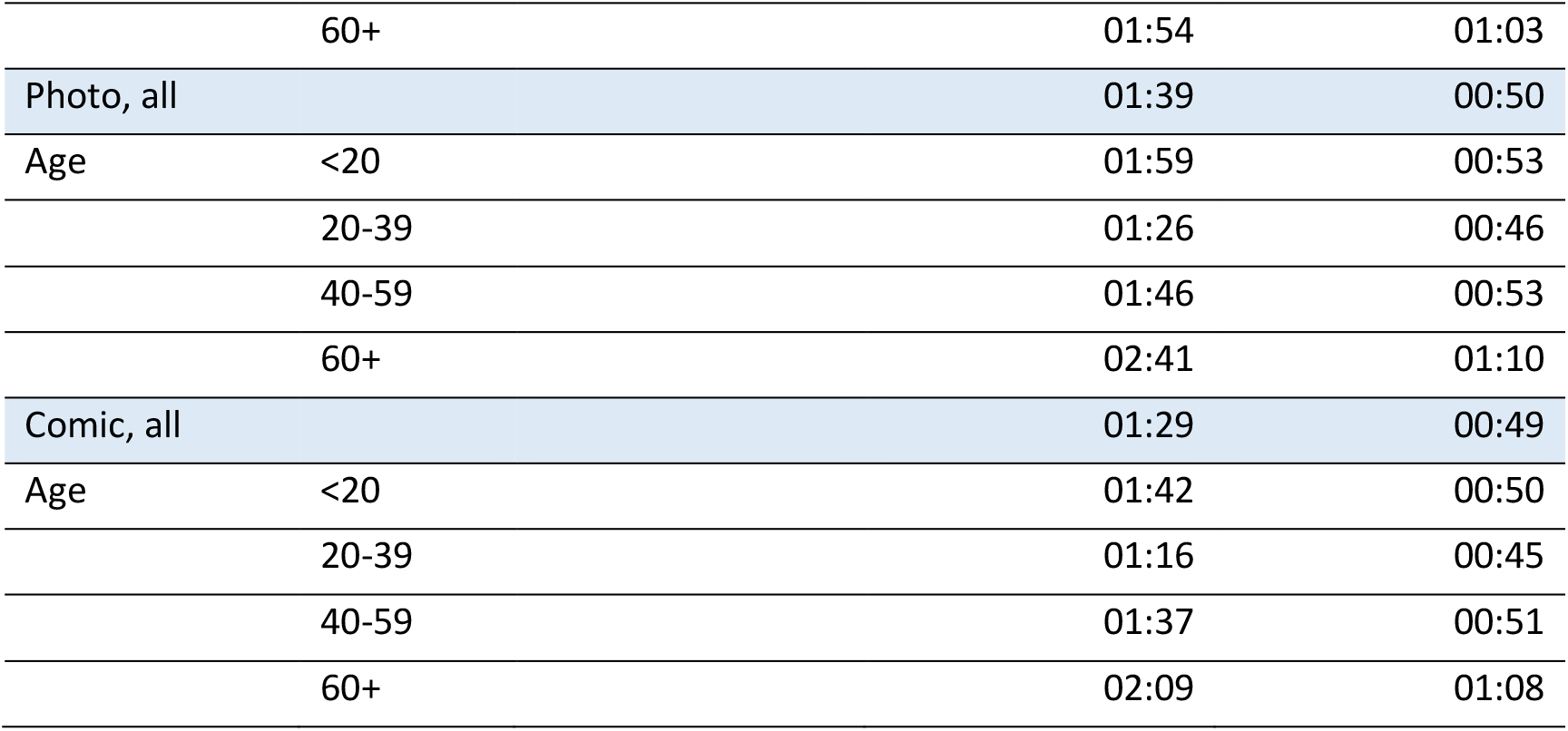
Summary of **Study 1**, pictogram survey. Median total response times (mimutes:seconds).

**Supplementary Table 3.** Question used in survey 2 and 3 and the information queried.

| Questions | Patient location | Technical information | Timing of Procedure | Frequency of Procedure |
| --- | --- | --- | --- | --- |
| **Study 2** Information delivery formats for cancer treatment timelines |  |  |  |  |
| Q1 - How often is a blood count taken during your first inpatient stay? |  |  | X | X |
| Q2 - When does the bone marrow puncture occur during the first inpatient stay? |  |  | X |  |
| Q3 - What happens between inpatient stays? | X |  |  |  |
| Q4 - When are you in the hospital? | X |  |  |  |
| Q5 - How long can the follow-up last? |  |  | X |  |
| Q6 - Where are you during the follow-up? | X |  |  |  |
| Q7 - How is the chemotherapy administered? |  | X |  |  |
| Q8 - How long is the chemotherapy administered? |  | X | X |  |
| Q9 - When is the chemotherapy administered? |  | X | X |  |
| Q10 - When does the stem cell transplantation take place? |  |  | X |  |
| **Study 3** Clinical evaluation of visual treatment timelines |  |  |  |  |
| Q1 - How long will you be in the clinic for treatment (at least)? | X |  | X |  |
| Q2 - When will you receive the [autologous blood stem cells/allogeneic blood stem cells/CAR-T cells]? |  |  | X |  |
| Q3 - How are the blood stem cells administered? |  | X |  |  |
| Q4 - How often do you come for follow-up care to the hospital? |  |  | X | X |
| Q5 - How do you receive the conditioning chemotherapy? |  | X |  |  |

**Supplementary Table 4.** Summary of **Study 2 – Comparing information delivery formats for cancer treatment paths**, 160 respondents.

| Group | Observation | mean | sd | min | max | range | se |
| --- | --- | --- | --- | --- | --- | --- | --- |
| Audio | Response quality | 0.67 | 0.17 | 0.3 | 1 | 0.7 | 0.02 |
|  | Confidence rating | 0.55 | 0.16 | 0 | 1 | 1 | 0.02 |
|  | Rating of info quality | 0.62 | 0.23 | 0 | 1 | 1 | 0.03 |
|  | Total response time | 14.23 | 5.76 | 6.59 | 41.13 | 34.54 | 0.74 |
|  | Total response time confidence | 4.19 | 0.9 | 2.8 | 7.36 | 4.56 | 0.12 |
| Pictogram | Response quality | 0.82 | 0.14 | 0.5 | 1 | 0.5 | 0.02 |
|  | Confidence rating | 0.78 | 0.13 | 0.5 | 1 | 0.5 | 0.02 |
|  | Rating of info quality | 0.79 | 0.19 | 0.33 | 1 | 0.67 | 0.03 |
|  | Total response time | 21.43 | 9.22 | 9.17 | 63.37 | 54.2 | 1.35 |
|  | Total response time confidence | 4.82 | 1.27 | 3.06 | 9.08 | 6.01 | 0.19 |
| Text | Response quality | 0.84 | 0.12 | 0.5 | 1 | 0.5 | 0.02 |
|  | Confidence rating | 0.82 | 0.11 | 0.6 | 1 | 0.4 | 0.01 |
|  | Rating of info quality | 0.76 | 0.21 | 0.33 | 1 | 0.67 | 0.03 |
|  | Total response time | 21.71 | 9.5 | 10.67 | 55.53 | 44.86 | 1.3 |
|  | Total response time confidence | 5.14 | 2.06 | 2.84 | 13.81 | 10.97 | 0.28 |

## Notes

### Competing Interest Statement

The authors have declared no competing interest.

### Funding Statement

The following fundings supported this study:
HKJ received a salary from a habilitation award of the Medical Faculty of the Technische Universitaet Dresden.
HKJ and MB received project funding from the Hochschulstiftung Medizin Dresden.
MB received funding from the MSNZ program of the Deutsche Krebshilfe.
The funders had no role in study design, data collection and analysis, decision to publish, or preparation of the manuscript.

### Author Declarations

The clinical evaluation, including the patients consent form, was approved by the TU Dresden ethics board (BO-EK-338072022).

### Summary of Updates

To improve clarity we have improved figures, tables, expanded the methods, changed the title, abstract and included further references.

